# Structural Racism, Neighborhood Opportunity, and Racial/Ethnic Disparities in Homicide Risk

**DOI:** 10.64898/2026.06.29.26356868

**Authors:** Robert W. Ressler, Manning Zhang, Madeline Leonardos, Dolores Acevedo-Garcia, Clemens Noelke

## Abstract

Homicide is a leading cause of preventable death in the United States and disproportionately affects Black and Indigenous communities. Structural racism and neighborhood disinvestment are central drivers of these disparities, yet national evidence on whether the association between neighborhood opportunity and homicide risk varies by race/ethnicity remains limited. Using 2020 data from the restricted-use National Violent Death Reporting System linked to Child Opportunity Index (COI 3.0) scores and Census population denominators across 30,077 ZIP codes in 48 states, we estimated age-adjusted log-linked generalized linear models to examine racial/ethnic disparities in homicide rates and their interaction with neighborhood opportunity. Black men experienced homicide rates nearly 20 times those of White men; Indigenous men experienced rates approximately 6 times higher. Higher neighborhood opportunity was independently associated with lower homicide risk across all groups and explained 43–59% of excess risk for Black and Indigenous individuals. Crucially, the association between neighborhood opportunity and homicide was nonlinear and significantly heterogeneous by race/ethnicity, with the steepest rate reductions occurring at the lowest opportunity levels for Black and Indigenous men. These findings suggest that place-based investments in severely deprived communities may yield the greatest reductions in homicide and racial/ethnic health inequities.

## Introduction

Homicide is a leading cause of preventable death and a significant public health crisis in the United States, disproportionately affecting Black and Indigenous communities (Martin, Rajan et al. 2022, Brosco and Payne 2023, Lanfear, Bucci et al. 2023, Krieger 2024, Zalla and Lesko 2024, Ranapurwala, Coles et al. 2025). According to CDC data, in 2020 the crude death rate for assault was 30.4 (per 100,000) for Black, 14.0 for Indigenous and 3.1 for White people. In addition, homicides play a key role in reducing life expectancy overall, and meaningfully contribute to racial/ethnic gaps in life expectancy (Light and Vachuska 2024).

Neighborhood opportunity is an important driver of homicide mortality, and racial/ethnic inequities in access to neighborhood opportunity likely explain a sizable part of the racial/ethnic inequities in homicide mortality (Peterson 2010, Sharkey and Faber 2014, Sampson, Wilson et al. 2018, Martin, Rajan et al. 2022, Blatt, Sadler et al. 2024, Randolph, Gonzalez-Guarda et al. 2024, Sullivan, Tian et al. 2024). However, whether the protective association between neighborhood opportunity and homicide rates varies across racial/ethnic groups is unclear. If the association is stronger among marginalized groups, that would suggest that investments in very low opportunity neighborhoods might be maximally efficient and likely reflect a pathway to narrowing persistent racial disparities (Cintron, Adler et al. 2022). To our knowledge, this is the first study leveraging national data to examine the independent and interactive effects of race/ethnicity and neighborhood opportunity on homicide mortality.

Studies using nationally representative data demonstrate that although homicide rates over the past two decades have declined for all racial/ethnic groups, between-group gaps remain significant (Light and Ulmer 2016). Average gun homicide rates for males in Black neighborhoods exceed those in white neighborhoods by a factor of fifteen to one, and structural disadvantages remain one of the strongest predictors of these disparities, serving as the primary pathway translating historical inequities into present day individual outcomes (Light and Ulmer 2016; Peterson and Krivo 2010, Sharkey and Faber 2014). More generally, children living in “very low” opportunity neighborhoods face a 30 to 40% higher risk of mortality, and historical policies explain nearly half the variation in contemporary childhood opportunity (Slopen, Cosgrove et al. 2023, Blatt, Sadler et al. 2024). Ultimately, these disparities are driven by a multidimensional array of neighborhood mechanisms that range from concentrated poverty to environmental stressors which collectively compromise the geography of opportunity that then shapes homicide risk (Acevedo-Garcia, Noelke et al. 2020).

Regarding the effect of neighborhood conditions on violence, prior research finds that structural disadvantage predicts homicide across Black, White, and Latino groups in the same direction, though effect magnitudes vary, which suggests that uniform structural effects across groups should not be assumed (Steffensmeier, Ulmer et al. 2010, Light and Ulmer 2016). When reassessing research on race, neighborhoods and violent crime, Sampson, Wilson, and Katz (2018) confirmed broad support for structural disadvantage as a cross-racial predictor of homicide, but also conclude that whether threshold or diminishing returns effects characterize the association between neighborhood disadvantage and homicide across racial groups remains untested.

Recent empirical studies have advanced our understanding of how racialized neighborhood conditions shape violence, but important gaps remain. Studies have been limited to cities and metropolitan areas, measured race or homicide mortality at the neighborhood-level, and relied on deprivation rather than opportunity measures (Light and Thomas 2019). Sullivan et. al. (2024) find that the Child Opportunity Index (COI; a neighborhood opportunity measure described below) partially mediated the relationship between census tract racial/ethnic predominance and violence-related mortality among children and adolescents, but their study was geographically limited to a single city, measured race/ethnicity at the neighborhood level and used a limited measure of violence-related mortality as an outcome. Gobaud et al. (2022) used data from 500 ZIP codes and a matched case-control design to show that lower ZIP-code household income has a sizeable effect on homicide rates, but the matching design only conditioned on racial/ethnic neighborhood composition and did not test for interaction in the association between race/ethnicity and ZIP code income. Cheon et al. (2020) used national data to show that the census tract proportion of Black residents and its level of socioeconomic deprivation are individually and interactively associated with gun homicide rates, with stronger associations between the proportion Black residents and homicide rates at higher levels of deprivation. But their work focuses on gun-related deaths rather than all homicides and measured racial/ethnic composition at the neighborhood level. Together, these studies show that racialized neighborhood conditions are strongly associated with violence, but do not test whether individual race/ethnicity modifies the associations between neighborhood opportunity and homicide rates across the U.S.

This study addresses these gaps by examining whether the association between neighborhood opportunity and homicide mortality varies by individual race/ethnicity. Using nationally representative data from the National Violent Death Reporting System (NVDRS) and the 2020 Decennial Census, we construct race/ethnicity-specific homicide rates at the neighborhood level rather than relying on neighborhood racial/ethnic composition as a proxy for racial/ethnic group-specific risks. This is possible because the NVDRS includes information both on decedent’s race and home ZIP code. Using the COI 3.0, we shift the framing and measurement of neighborhood conditions from a focus on deficits to a focus on multidimensional assets and opportunities. Estimating the interaction between race/ethnicity and neighborhood opportunity is important because average neighborhood effects may obscure whether opportunity is most strongly associated with reduced homicide risk among the groups and places experiencing the greatest burden. Identifying such heterogeneity can clarify whether place-based investments in very low-opportunity neighborhoods are likely to reduce homicide overall, narrow racial/ethnic inequities, or both. We hypothesized that higher neighborhood opportunity would be associated with reduced homicide risk among all groups, but that this association would be larger among Indigenous and Black individuals, reflecting both elevated homicide rates relative to White or Hispanic Americans and segregation into low opportunity neighborhoods.

## Methods

### Study Design, Population, and Settings

The analytic dataset was constructed by integrating three sources: the NVDRS restricted-use file, ZIP Code Tabulation Area (ZCTA)-level population counts from the 2020 Decennial Census, and ZIP code-level COI 3.0 scores.

The NVDRS file was subset to incident year 2020. To account for incomplete county-level reporting in the NVDRS for California and Texas, we constructed a geographic inclusion variable and retained ZCTAs only if at least 90% of their population resided within a reporting county. This alignment ensures that homicide counts and denominators reflect the same underlying geography. Following the total exclusion of Florida and Hawaii for non-reporting status, the final analytic sample was reduced from 331,449,281 individuals (Census 2020 population estimate) to 278,483,236, representing 85% of the total population. Demographically, the sample remains highly representative, containing 2.1% fewer Hispanic individuals and maintaining all other racial and ethnic categories within 1% of national estimates. Regarding neighborhood distribution, the analytic sample contains 3.1% fewer low-opportunity and 1.9% more high-opportunity neighborhoods than the national average.

The NVDRS contains the following manners of death: homicide, suicide, unintentional firearm death, legal intervention, and undetermined intent. We focus on the homicide category, which counts all homicides including intentional gun deaths. Continuous age in years was grouped into five-year intervals (0–4, 5–9, …, 85+) to align with Decennial Census data. Race/ethnicity was harmonized across the NVDRS and Census files into five analytic categories: White non-Hispanic (White), Black non-Hispanic (Black), Hispanic, American Indian/Alaska Native non-Hispanic (Indigenous), and Asian/Pacific Islander non-Hispanic (Asian); the latter combined Asian and Native Hawaiian/Other Pacific Islander subgroups in both data sources to ensure comparability. Those with more than one race were categorized as Two Plus, and all others as Other. We excluded records with missing values on age, sex, race/ethnicity, decedent’s home ZIP code, or death state. The final cleaned NVDRS file contained 19,960 homicides, representing 94% of the 21,195 homicides in the analytic states and 81% of the CDC-reported 24,535 homicides nationally in 2020.

NVDRS homicide counts were collapsed to the ZIP code-by-race/ethnicity-by-sex-by-age-groups level. The resulting count file was merged with the ZCTA population denominators from the 2020 Decennial Census, so that ZCTAs with zero observed homicides were retained and their homicide counts set to zero. COI 3.0 Child Opportunity Scores were merged onto the analysis file using ZIP codes.

### Dependent Variable

Homicide rates per 100,000 persons were computed within each ZCTA-race/ethnicity-sex-age stratum as 100,000 × (*h*/*n*), where *h* is the observed homicide count and *n* is the stratum-specific Census population denominator.

### Neighborhood Opportunity

We measured neighborhood opportunity at the ZIP code-level using the Child Opportunity Index (COI) 3.0, which ranks U.S. Census tracts from 1 to 100 based on 44 indicators covering three domains: education, health and environment, and socioeconomic resources. The COI 3.0 is a multidimensional measure incorporating features criminologists link to homicide rates, such as economic deprivation, vacant homes, and heat exposure (Branas, South et al. 2018, Kondo, Andreyeva et al. 2018, Moyer, MacDonald et al. 2019, South, MacDonald et al. 2023, Choi, Heo et al. 2024). Unlike the Social Vulnerability Index, the COI does not include measures of neighborhood racial/ethnic composition or language in its construction, it allows neighborhood opportunity to be modeled separately from individual race/ethnicity and from the racialized sorting of populations across neighborhoods (Acevedo-Garcia, Noelke et al. 2024, Noelke, McArdle et al. 2025).

### Statistical Analysis

To evaluate the independent and joint associations of race/ethnicity and neighborhood opportunity with homicides, we estimated generalized linear models (GLMs) with a log link separately for males and females. We explored different approaches to model the curvilinear association between COI and homicide mortality observed in the raw data. The GLM with a log link fit the curvilinear relationship well. All models were weighted by stratum population size and adjusted for age group, with White (non-Hispanic) as the reference racial/ethnic category and the 45–49 age group as the reference age category. Standard errors were clustered at the ZCTA/ZIP code level throughout to account for the non-independence of observations within the same neighborhood.

We ran the following models: Model 1 included race/ethnicity indicators and age to quantify age-controlled racial/ethnic disparities in homicide risk. Model 2 included the continuous COI national percentile score and age group, with no race/ethnicity terms, to estimate the age-controlled association between neighborhood opportunity and homicide rate. Model 3 adds back group-level indicators for race/ethnicity along with the COI score and Model 4 adds interactions between race/ethnicity indicators and COI, allowing the slope of the opportunity–homicide relationship to differ across racial/ethnic groups. The joint significance of the interaction terms was evaluated using a Wald test. Predicted marginal homicide rates from Model 4 were estimated at each integer value of the COI score (1–100) for each racial/ethnic group. We present exponentiated coefficients (incidence rate ratios [IRRs]) and 95% confidence intervals (CIs), which are interpreted as the multiplicative change in the outcome relative to the reference category.

This study was reviewed by Boston University’s Institutional Review Board (Protocol #7997X) and determined to be exempt from full review. All data linkages and analyses were performed using Stata 18. Large Language Models (OpenAI’s ChatGPT and Anthropic’s Claude) were used in the production of this manuscript to support coding, description of methods, drafting and revisions of the manuscript and abstract.

As a sensitivity analysis, we assessed whether the association between neighborhood opportunity and homicide risk varied by urban-rural classification. ZIP codes were classified as urban (RUCA codes 1–3) or rural (RUCA codes 4–10) using the USDA Rural-Urban Commuting Area (RUCA) codes. The three main model sequences (race-only, COI-only, and race + COI) were re-estimated separately for urban and rural ZIP codes. To formally test whether urban-rural status moderated the COI-homicide association, we estimated an interaction model including race/ethnicity, COI, urban-rural status, and a COI × urban-rural interaction term, with joint significance evaluated using a Wald test.

## Results

### Study Population

Table 1 describes the demographic and neighborhood characteristics covering 136,625,658 men and 141,857,578 women, representing 85.06% of the population of the United States in 2020. Crude homicide rates in our analytic sample closely correspond to published CDC WONDER estimates for 2020. For example, our rate for non-Hispanic Black men (60.93 per 100,000) is only 0.73 above the CDC upper confidence bound of 60.2 for that group. Although the rate for White men (3.91) is below the CDC lower confidence bound of 4.2, our rates for non-Hispanic Black women (8.46), Indigenous men (21.76) and Indigenous women (6.51) each fall within to their respective CDC 95% confidence intervals, confirming that our sample is highly representative of the national homicide burden across racial/ethnic groups. Descriptively, Black men experienced a homicide rate 57.02 per 100,000 higher than White men (60.93 vs 3.91) and Indigenous men a rate 17.85 per 100,000 higher (21.76 vs 3.91). For Black and Indigenous women, the absolute rate differences compared to White women were 6.60 (8.46 vs 1.86) and 4.65 (6.51 vs 1.86) per 100,000, respectively. Across racial/ethnic groups, individuals from high-opportunity neighborhoods are 4-7 times less likely to die by homicide than those from low-opportunity neighborhoods. Notably, the lowest Black homicide rate found in very-high opportunity neighborhoods (11.14 per 100,000) is still larger than the highest White homicide rate in very-low opportunity neighborhoods (7.44 per 100,000).

**Table 1.** Descriptive Statistics from the NVDRS, Census, and COI.

|  | COI 3.0 Opportunity Levels |  |  |  |  |  |  |
| --- | --- | --- | --- | --- | --- | --- | --- |
|  | Men | Women | Very Low | Low | Moderate | High | Very High |
| Black |  |  |  |  |  |  |  |
| Homicide Count | 10003 | 1539 | 6599 | 2417 | 1339 | 839 | 348 |
| Population | 16418241 | 18190218 | 13108927 | 7903503 | 5791450 | 4681238 | 3123341 |
| Homicide Rate | 60.93 | 8.46 | 50.34 | 30.58 | 23.12 | 17.92 | 11.14 |
| White |  |  |  |  |  |  |  |
| Homicide Count | 3245 | 1581 | 1162 | 1399 | 1006 | 816 | 443 |
| Population | 82936161 | 85170749 | 15610421 | 31393033 | 36738117 | 41459604 | 42905735 |
| Homicide Rate | 3.91 | 1.86 | 7.44 | 4.46 | 2.74 | 1.97 | 1.03 |
| Hispanic |  |  |  |  |  |  |  |
| Homicide Count | 2369 | 456 | 1479 | 595 | 380 | 240 | 131 |
| Population | 22358848 | 22417772 | 14375101 | 9505737 | 8300279 | 7040089 | 5555414 |
| Homicide Rate | 10.60 | 2.03 | 10.29 | 6.26 | 4.58 | 3.41 | 2.36 |
| Asian |  |  |  |  |  |  |  |
| Homicide Count | 167 | 67 | 36 | 58 | 52 | 46 | 42 |
| Population | 7640877 | 8427692 | 1298936 | 2129201 | 2699840 | 3543199 | 6397393 |
| Homicide Rate | 2.19 | 0.79 | 2.77 | 2.72 | 1.93 | 1.30 | 0.66 |
| Indigenous |  |  |  |  |  |  |  |
| Homicide Count | 224 | 69 | 184 | 46 | 43 | -- | -- |
| Population | 1029241 | 1060525 | 927618 | 433418 | 351005 | 264926 | 112799 |
| Homicide Rate | 21.76 | 6.51 | 19.84 | 10.61 | 12.25 | -- | -- |
| Two or More Races |  |  |  |  |  |  |  |
| Homicide Count | 162 | 48 | 61 | 66 | 35 | 32 | 16* |
| Population | 5528386 | 5888006 | 1526434 | 2239691 | 2428688 | 2625454 | 2596125 |
| Homicide Rate | 2.93 | 0.82 | 4.00 | 2.95 | 1.44 | 1.22 | 0.62 |
| Other |  |  |  |  |  |  |  |
| Homicide Count | 30 | 13* | -- | 13* | -- | -- | -- |
| Population | 713904 | 702616 | 215035 | 264599 | 312983 | 299009 | 324894 |
| Homicide Rate | 4.20 | 1.85* | -- | 4.91* | -- | -- | -- |

-- = Suppressed (count <10)
\* = Unstable estimate (count <20); interpret with caution

### Regression

Log-linked age-controlled generalized linear models estimated death rates for 30,084 unique ZIP codes for men and 30,079 for women. Table 2 presents adjusted average marginal effects (AMEs) from age-adjusted log-linked GLMs estimated separately for men and women. Race/ethnicity and neighborhood opportunity are both significant predictors of homicide rates.

**Table 2.**
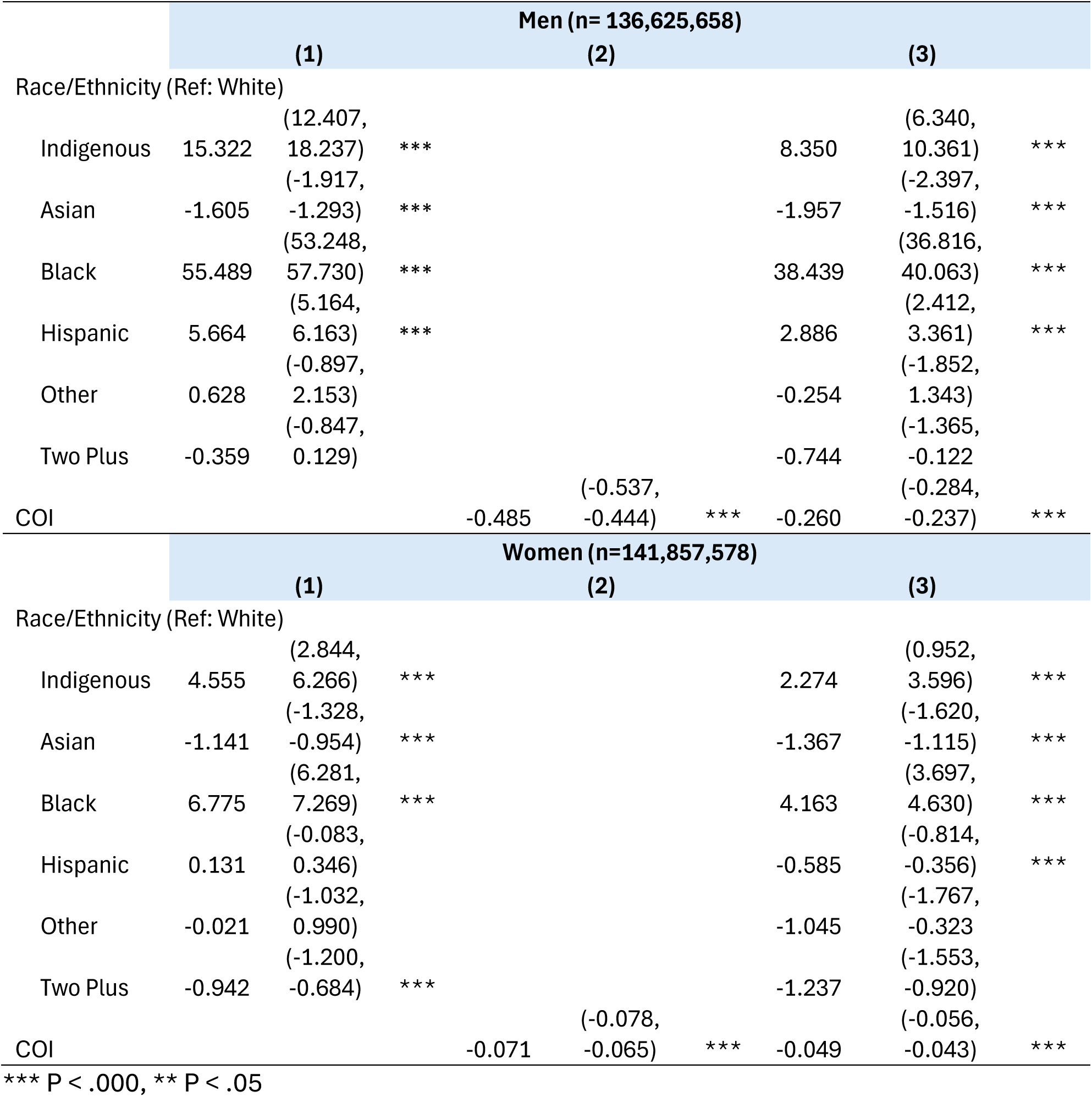
Average Marginal Effects from Age-Controlled Log-Linked Generalized Linear Models for Men and Women Predicting Homicide Rate with Race/Ethnicity and COI.

Model 1 (race/ethnicity only) confirms large and statistically significant racial/ethnic disparities relative to White individuals that reproduce the descriptive results in Table 1. Black men had an age-adjusted homicide rate of 55.49 additional deaths per 100,000 higher than White men (95% CI: 53.25, 57.73), and Indigenous men had a rate of 15.32 per 100,000 higher (95% CI: 12.41, 18.24). Among women, the corresponding AMEs were 6.78 (95% CI: 6.28, 7.27) for Black women and 4.56 (95% CI: 2.84, 6.27) for Indigenous women. Asian men and women had significantly lower rates than White peers (AME: −1.61 and −1.14 per 100,000, respectively). The Hispanic coefficient for women was not statistically significant in Model 1 (AME: 0.13; 95% CI: −0.08, 0.35). The Other and Two Plus categories are not significantly different from the White reference group for men, and Two Plus is significant for Women (-0.942, p < .000), but we caution against interpreting these results because of the ambiguity of these race/ethnicity categories.

Model 2 (COI only) shows that neighborhood opportunity alone is inversely associated with homicide risk. Each one-point increase in the COI national percentile score was associated with a decrease of 0.49 deaths per 100,000 for men (95% CI: −0.54, −0.44) and 0.07 per 100,000 for women (95% CI: −0.08, −0.07). These non-overlapping confidence intervals suggest that the protective association of neighborhood opportunity with homicide rates was far stronger among men than women.

In sensitivity analyses stratifying by urban-rural classification, the association between neighborhood opportunity and homicide risk was consistent across settlement types. A formal test of the COI × urban-rural interaction was not statistically significant for men (chi2 = 1.67, P = .197) or women (chi2 = 0.03, P = .874), indicating that the inverse relationship between neighborhood opportunity and homicide risk does not differ meaningfully between urban and rural areas (detailed results available on request).

Model 3 indicates that race/ethnicity and neighborhood opportunity are each independently associated with homicide rates after mutual adjustment. Introducing COI into the race/ethnicity model substantially attenuates but does not eliminate racial/ethnic disparities. For men, the AME for Black men relative to White men decreased from 55.49 to 38.44 per 100,000, (a 31% decrease) and the AME for Indigenous men decreased from 15.32 to 8.35 per 100,000 (46%). For women, the AME attenuated from 6.78 to 4.16 for Black women (a 39% decrease) and from 4.56 to 2.27 for Indigenous women (50%). Conversely, adding race/ethnicity to the COI model reduced the magnitude of the opportunity coefficient: the per-point AME of COI decreased from −0.49 to −0.26 per 100,000 for men and from −0.07 to −0.05 for women (a decrease of 46% and 31%, respectively). Notably, for Hispanic women, the introduction of COI reversed the direction of the coefficient and rendered it statistically significant (AME: 0.13 to −0.59), indicating that after accounting for neighborhood opportunity, Hispanic women have a significantly lower homicide rate than White women (P < 0.001). Other and Two Plus are insignificantly different from the White reference in these models.

Figure 1 displays the predicted homicide rates over COI scores for each of the racial/ethnic groups in the analysis from a model which includes an interaction term between race/ethnicity and COI (model not shown but available upon request, Wald test for the interaction term: chi2=2106 for Men, 246 for Women, both P < 0.000). The interaction terms (e.g. slope) for Black, Hispanic, Asian, and Other men x COI are significantly different from White men (P < 0.05) and for Black, Hispanic, and Other women compared to White women (P < 0.05). For Indigenous men, and particularly for Black men, increases in COI scores are associated with greater reductions in homicide rates at lower ends of the opportunity distribution, leading to an especially steep downward slope within the bottom 20% or very low-opportunity category. For clarity we opted not to include Other and Two Plus in these visualizations, but in results not shown predicted rates for the Two Plus and Other groups among men closely track those of White men across the opportunity distribution. Among women, these two groups exhibit some of the lowest predicted homicide rates, falling below White and Hispanic women particularly at the lowest levels of neighborhood opportunity.

**Figure 1:**
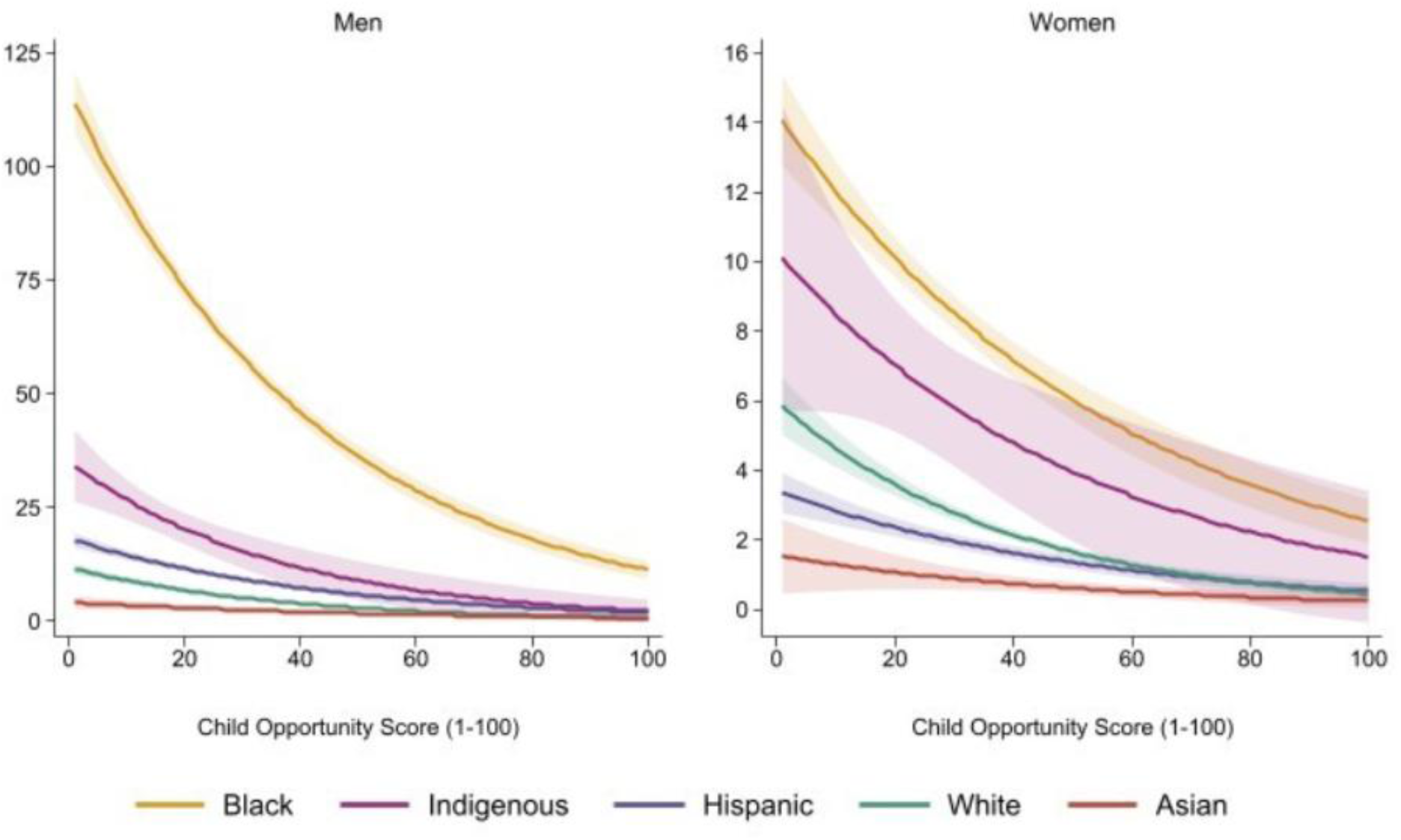
Predicted Homicide Rate by COI Score, Race/Ethnicity, and Gender

## Discussion

Prior studies of racialized neighborhood conditions and violent deaths have been confined to single cities or metropolitan areas or have used neighborhood racial/ethnic composition as a proxy for group-specific risk (Cheon, Lin et al. 2020, Gobaud, Mehranbod et al. 2022, Sullivan, Tian et al. 2024). Our study uses 2020 homicide data with victim-level information on race/ethnicity and home ZIP codes, allowing us to separately quantify the association between race/ethnicity, neighborhood opportunity and homicide rates. To our knowledge, this is the first national study to show how the association between neighborhood opportunity and homicide varies across (male and female) Black, White, Hispanic, Asian, and Indigenous populations. We found substantial racial/ethnic inequities in homicide that persisted after adjustment for neighborhood opportunity, alongside an inverse association between neighborhood opportunity and homicide that was substantially stronger among the groups bearing the greatest burden, Black and Indigenous men. After age adjustment, Black and Indigenous men experienced 55.5 and 15.3 additional homicide deaths per 100,000, respectively, relative to White men; the corresponding differences among women were 6.8 and 4.6 per 100,000. Adjustment for neighborhood opportunity attenuated the Black–White and Indigenous–White disparities by roughly one-third to one-half. The association between neighborhood opportunity and homicide also varied significantly by race/ethnicity. Among both Black and Indigenous men, the decline in predicted homicide rates was concentrated at the low end of the opportunity distribution. Black male (Indigenous male) homicide mortality differed by 102.80 deaths per 100,000 (31.94 per 100,000) between neighborhoods with the lowest opportunity (COI score of 1) and the highest opportunity (COI score of 100), while the difference for White males was 10.74 deaths per 100,000. Lastly, across the neighborhood opportunity distribution, substantial Black-White differences persisted. In very high opportunity neighborhoods, the homicide rate among Black men (11.14 per 100,000) exceeded that among White men (1.03 per 100,000) and also exceeded homicide mortality among White men in very low opportunity neighborhoods (7.44 per 100,000).

The significant interaction between COI and race/ethnicity in our models has two distinct but not mutually exclusive explanations. The first is that structural neighborhood factors unmeasured in the COI and specific to neighborhoods where Black and Indigenous people reside are resulting in stronger associations between COI and homicide mortality for these groups at the low end of the opportunity distribution. The second is that neighborhood opportunity is related to homicide through qualitatively different mechanisms for different racial/ethnic groups. In either case, the findings underscore the central role of socio-structural determinants in shaping homicide risk across all groups, the role of historical structural racism in particular.

First, the COI does not capture all the possible neighborhood mechanisms that might influence experiences of violence, and which are mechanisms that are specific to minoritized groups. For example, measures are still limited regarding safety, over-policing and the prison industrial complex, forced displacement and the reservation system, and the compounding effects of persistent and generational marginalization. These factors contribute to the embodiment of structural disadvantage in ways that are omitted in measures that focus on immediate neighborhood conditions (Geronimus, Hicken et al. 2006). Nevertheless, considering structural racism as an underlying cause of disparities in health means that these measures outside of the COI would still be expected to vary systematically by race/ethnicity and could be driving the significant interactions (Phelan and Link 2015).

Second, the COI does measure multiple pathways through which the structural environment could be expected to influence homicide violence differently for different groups. In this discussion, we focus on three mechanisms we believe most predict variation by racial/ethnic identity and therefore contribute to explaining the observed results: Institutions, Poverty, and Collective Efficacy.

### Pathways Linking Race/Ethnicity, Neighborhood Opportunity, and Violence

This analysis refines our understanding regarding variation in the pathways connecting structural conditions to violence by racial or ethnic group. Conceptually, we contribute to this literature by situating our analysis within an opportunity rather than deprivation framework, and by including Indigenous, Hispanic, and Asian groups. Methodologically, analyzing all homicides with data from nearly the entire United States and accounting for nonlinearity adds depth and context to research on effect heterogeneity in violence by race/ethnicity. Both Black and Indigenous communities experience disproportionately high rates of violence that are fundamentally rooted in structural conditions like concentrated poverty, racial segregation, historical oppression, and institutional disinvestment. These structural disadvantages are qualitatively different from those experienced by White, Asian, and Hispanic groups, and they produce compounding effects and further mediate the risk of violence.

#### Institutions

Neighborhood institutions like schools, clinics, and community organizations play a critical role in shaping health outcomes. However, these institutions are not immune to the effects of structural racism, which produces racially disparate outcomes regardless of individual intent or overt discrimination (Bonilla-Silva 2021). When institutions are located within or adjacent to low-opportunity neighborhoods, their ability to meet the specific needs of the racial/ethnic majority in that area may be enhanced (Randolph, Gonzalez-Guarda et al. 2024). These higher returns to the same services could help explain why Black and Indigenous individuals appear to benefit more from increases in neighborhood opportunity at the lower end of the distribution.

#### Poverty

Poverty is a well-established contributor to violence (Cheon et al. 2020). Yet for many racial and ethnic minority communities, poverty is not solely economic. It is structural, multigenerational, and compounded by systemic exclusion (Phelan and Link 2015). Neighborhoods with high concentrations of minoritized populations often experience amplified effects of poverty due to limited access to mobility-enhancing resources(Kravitz-Wirtz, Bruns et al. 2022). Under these conditions, improvements in neighborhood opportunity may yield disproportionately high marginal returns, particularly in low-opportunity areas.

#### Collective Efficacy

The ability of residents to form strong social bonds and act collectively to prevent harm is known as collective efficacy (Sampson, Raudenbush et al. 1997). It is another key pathway through which neighborhood opportunity influences violence (Sharkey and Faber 2014). The capacity for collective efficacy is shaped not only by material conditions, but also by the historical relationship between communities and structural racism (Mueller-Williams, Coughlin et al. 2024). For Black and Indigenous communities, these resources can foster social cohesion and resilience, and counter the isolating effects of structural violence. Access to local cultural institutions, parks, and public gathering spaces, features captured in the COI, may be especially important for communities that have historically been denied safe public spaces to gather.

Regardless of the driving force underlying the observed significant interaction, neighborhood opportunity is not merely about access to amenities, it is a structural determinant of violence. These findings highlight that differences in health outcomes across racial and ethnic groups cannot be explained solely by individual behaviors or immediate circumstances; instead, they reflect long-standing structural factors and historical conditions that systematically place marginalized communities at higher risk. Although it is sometimes hypothesized that the association between violence and ever-increasing disadvantage might level off at the extremes, our analysis indicates that even small improvements in neighborhood opportunity at the low end of the distribution could spur meaningful reductions in homicides. Our findings therefore underscore the urgent need for place-based strategies that expand opportunity and redress the cumulative harms of structural racism.

### Strengths and Limitations

This study offers several key strengths. First, it draws on national data, enabling a comprehensive view of neighborhood opportunity and violence across diverse geographic and demographic contexts. Second, by focusing specifically on homicides, the analysis centers on the most severe and measurable form of interpersonal violence. Third, the inclusion of victim race/ethnicity allows for an intersectional approach that examines how structural conditions shape risk differently across racialized groups (Peterson 2010).

Several limitations should be acknowledged. First, the data reflect patterns from 2020—a year marked by heightened violence due to the COVID-19 pandemic and social unrest.

Although 2020 saw a 24% increase in homicides, these excess deaths did not track with COVID-19 infection waves, suggesting they were driven by underlying social and structural vulnerabilities (Zalla and Lesko 2024). This justifies the use of the COI 3.0 to examine how pre-existing inequities in neighborhood opportunity modulated homicide risk during a period of acute social stress and while this context may limit generalizability, it also provides insight into structural dynamics under duress. Second, the analysis uses the victim’s residence rather than the location of the violent incident. While this aligns with the study’s focus on neighborhood opportunity, it may not fully capture the situational context of violence. Future research could incorporate incident location data to assess how often violence occurs outside the home neighborhood and whether these patterns vary by race, age, or gender. Third, the race and ethnicity categories used are necessarily broad, which may obscure important within-group variation. Additionally, census data collection among Indigenous populations is subject to known inaccuracies, limiting precision. Although similar to national estimates in CDC WONDER, the exclusion of non-reporting counties in California and Texas may introduce misclassification bias or measurement error for the Hispanic estimates. More granular data—particularly for Indigenous, Hispanic, and Asian American populations—would allow for a more nuanced understanding of how neighborhood opportunity intersects with racialized experiences of violence. The COI does not capture all neighborhood-level mechanisms that influence violence, such as policing. It is possible that adding these to the model would attenuate racial/ethnic variation in the observed neighborhood effect. Finally, the analysis is associational rather than causal. Future studies should explore quasi-experimental or longitudinal designs to better establish causal pathways.

### Policy Implications

Our findings point to several actionable policy implications. Without including identity groups within its construction, The Child Opportunity Index offers a practical tool for identifying neighborhoods where investments could yield the greatest reductions in violence and racial/ethnic inequities. Furthermore, these neighborhood investments still need to proactively consider the specific racial and ethnic makeup of the neighborhood to determine best strategies for improving opportunity. These approaches move from asking “Do neighborhoods matter” towards understanding how, when, and for whom they matter, emphasizing the role of governments and inclusive community organizing in creating environments where safety and well-being can flourish for everyone (Sharkey and Faber 2014).

### Conclusion

Our findings demonstrate that neighborhood opportunity is a structural determinant of violence and that exposure to violence runs through institutions like schools, clinics, and public spaces. Violence must therefore be addressed as a racialized, multigenerational, and structural condition. Only then can interventions begin to dismantle the systemic barriers that perpetuate violence. These associations are not incidental; they reflect the mechanisms through which structural racism has historically denied safety and opportunity to Black, Indigenous, and other marginalized communities. Investing in neighborhood opportunity is therefore a viable strategy for achieving long-lasting reductions in racial/ethnic homicide disparities, public health advancement, and racial equity.

## Data Availability Statement

The NVDRS restricted-use data are available through application to the CDC (https://www.cdc.gov/nvdrs). COI 3.0 data are publicly available at diversitydatakids.org. Analysis code is available from the corresponding author upon request.

## Acknowledgements

We would like to thank Nancy McArdle, Yang Lu, and Brian DeVoe for their feedback on the development of this work, and Jon Foerschler for his support with the secure data computer room.

